# Cognitive functioning in anxiety and depression: Results from the ALSPAC cohort

**DOI:** 10.1101/2021.08.25.21262634

**Authors:** Steph Suddell, Liam Mahedy, Caroline Skirrow, Ian S. Penton-Voak, Marcus R. Munafò, Robyn E. Wootton

**Affiliations:** University of Bristol, School of Psychological Science, University of Bristol, UK; National Institute for Health Research Biomedical Research Centre at the University Hospitals Bristol NHS Foundation Trust and the University of Bristol, UK; MRC Integrative Epidemiological Unit, University of Bristol, UK; Cambridge Cognition, Cambridge, UK; Population Health Sciences, Bristol Medical School, University of Bristol, Bristol, UK; Nic Waals Institute, Lovisenberg Diaconal Hospital, Oslo, Norway

**Author notes:** **Corresponding author:** Steph Suddell, School of Psychological Science, University of Bristol, 12a Priory Road, Bristol, UK, BS8 1TU.

**Keywords:** Anxiety, Depression, Emotion Recognition, Working Memory, Cognition, Response Inhibition, ALSPAC

## Abstract

**Background:** Understanding the nature of cognitive deficits in anxiety and depression may identify intervention targets and help prevent functional decline. This study used observational and genetic methods to investigate the association of anxiety and depression with emotion recognition, response inhibition, and working memory, in young adulthood.

**Methods:** The Avon Longitudinal Study of Parents and Children (ALSPAC) is a large prospective birth cohort study. Participants completed regular postal questionnaires and in-clinic assessments, starting from September 6, 1990. Data collection is ongoing. Linear regression was used to assess 1) cross-sectional associations between anxiety, depression, and cognition at age 24 (*n* = 2,187) and 2) prospective associations between anxiety and depression at age 18 and cognition at age 24 (*n* = 1,855). Mendelian randomization analyses were conducted to assess causal pathways between anxiety, depression, and cognition.

**Results:** Primary analyses were conducted on 3,087 participants following multiple imputation. There was evidence for anxiety being associated with a decreased recognition of happiness (*b = -*0.27, 95% CI -0.54 to 0.01, *p* = .045), and depression being associated with an increased recognition of sadness (*b =* 0.35, 95% CI 0.07 to 0.64, *p* = .016). Anxiety was negatively associated with working memory (*b = -*0.14, 95% CI -0.24 to 0.04, *p* = .005), but no association was found for depression (*b =* 0.06, 95% CI -0.05 to 0.16, *p* = .284). There was no evidence for any association with response inhibition. Results from Mendelian randomization analyses were inconclusive, likely due to low statistical power.

**Conclusions:** There was little evidence that anxiety and depression are associated with significant impairments in executive functioning. However, both anxiety and depression were associated with altered emotion recognition. This may inform the development of interventions that target psychosocial functioning.

---

Anxiety and depression are leading causes of disability worldwide (World Health Organization, 2017). The conditions are highly comorbid (Essau et al., 2018), with over 50% of depressed individuals also having an anxiety disorder (Hirschfeld, 2001). Onset is common in adolescence and young adulthood, with earlier onset being associated with more severe trajectories (Le Roux et al., 2005; Park et al., 2014). In addition to the core symptoms of low mood and psychological distress, these conditions are often associated with poorer cognitive and psychosocial functioning (Knight et al., 2018) that persists even in remitted patients (Rock et al., 2014). Understanding the nature of these deficits may identify targets for intervention and help prevent further functional decline.

To date, much of this research has been conducted in relatively small-scale psychological studies with heterogenous designs, limiting comparisons across studies. Additionally, despite anxiety and depression being highly comorbid (Essau et al., 2018), their relationships with cognitive functioning are often studied in isolation. Meta-analyses synthesising this work suggest depressed individuals display moderate deficits in several domains of executive functioning, including attention, memory and processing speed (Lee et al., 2012; Semkovska et al., 2019; Snyder, 2013). Depressed individuals also show deficits in socio-cognitive domains, including impaired recognition of emotional facial expressions (Dalili et al., 2015). While fewer meta-analyses have been conducted on cognition in anxiety disorders, those available report broadly similar results, including deficits in working memory, learning, and emotional processing (Moran, 2016; O’Toole et al., 2013; Schuitevoerder et al., 2013; Shin et al., 2014).

As noted by de Nooij and colleagues (2020), meta-analyses may overinflate effect sizes if the included studies rely on samples not reflective of a general population, highlighting the need for population-based studies. Their investigation of the UK Biobank (aged 45-81 years) identified deficits in several domains of cognition, including executive functioning and processing speed associated with lifetime depression (De Nooij et al., 2020). However, effect sizes were smaller than in traditional case-control studies. Earlier work, also in the UK Biobank (aged 40-69 years), found evidence for cognitive deficits associated with lifetime recurrent depression in unadjusted analyses (Cullen et al., 2015). In contrast, research in the Generation Scotland study (average age 51 years; Meijsen et al., 2018) reported lower processing speeds in depressed participants, in addition to superior vocabulary scores. Although these studies shed light on the relationship between mental health and cognition in mid-to-late adulthood, there is a need to examine the relationship in young adulthood, when many cognitive functions reach maturity (Hartshorne & Germine, 2015) and emotional disorders often emerge (Le Roux et al., 2005; Park et al., 2014).

The Avon Longitudinal Study of Parents and Children (ALSPAC) is a large birth cohort that presents an opportunity to study the relationship between cognition and mental health in adolescence and young adulthood. In the present study, we examined the association between anxiety and depression with three domains of cognitive functioning in young adulthood: working memory, emotion recognition, and response inhibition. We aimed to (i) examine the cross-sectional association between anxiety and depression and cognition at age 24, (ii) conduct prospective analyses to explore the relationship between anxiety and depression at age 18 and cognition at age 24, and (iii) triangulate this observational work with genetic analyses, using Mendelian randomization to support stronger causal inference.

## Method

### Participants

ALSPAC recruited pregnant women residing in the former county of Avon, UK, who were due to give birth between 1 April 1991 to 31 December 1992. An initial 14,541 pregnancies were enrolled, leading to 14,062 live births and 13,988 children being alive at 1 year. Data was collected at regular intervals via postal questionnaires and in-clinic assessments. The study population that completed the cognitive assessments at age 24 (used in this study) is detailed in previous work (Mahedy et al., 2021). The study website contains details of all the data that is available through a fully searchable data dictionary and variable search tool (www.bristol.ac.uk/alspac).

Ethics approval for the study was obtained from the ALSPAC Ethics and Law Committee and the Local Research Ethics Committees. Written informed consent was obtained for the use of data collected via questionnaires and clinics from parents and participants following recommendations of the ALSPAC Ethics and Law Committee at the time. Consent for biological samples has been collected in accordance with the Human Tissue Act (2004). More details on ethics committees/institutional review boards are provided here: http://www.bristol.ac.uk/alspac/researchers/research-ethics/

### Measures

A timeline of variables can be seen in Figure 1. Study data were collected and managed using REDCap (Research Electronic Data Capture) which is a secure, web-based software platform supporting data capture in research hosted at the University of Bristol (Harris et al., 2009).

**Figure 1.**
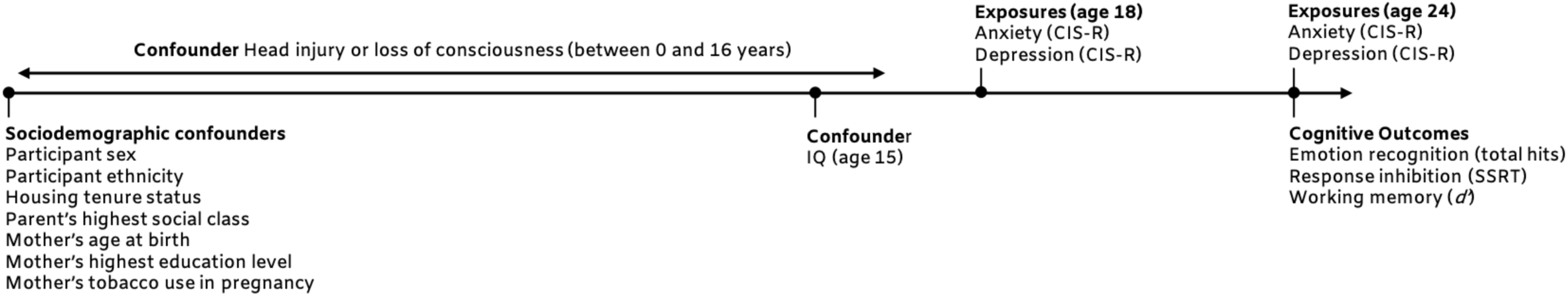
Timeline of variables used in the prospective and cross-sectional observational analyses. The outcome variables for all analyses were emotion recognition total hits (number of correctly identified facial expressions out of 96), response inhibition (as measured by SSRT = stop signal reaction time) and working memory (d’ = d prime, a discriminability index on the n-back task) at age 24. Exposures were anxiety and depression at ages 18 and 24, as measured by the Clinical Interview Schedule (Revised; CIS-R).

#### Anxiety and Depression

Anxiety and depression were assessed using the Revised Clinical Interview Schedule (CIS-R; Lewis et al., 1992) at both ages 18 and 24. The CIS-R is a self-administered computerized questionnaire which assesses a range of neurotic symptoms to derive suggested diagnoses based on ICD-10 criteria for anxious and depressive disorders. Two binary variables (present/absent) for anxiety and depression were derived. The anxiety variable captured all individuals with a primary or secondary diagnosis of generalized anxiety disorder, social phobia, agoraphobia, panic, and non-specific neurotic disorders. The depression variable captured all individuals with a primary or secondary diagnosis of mild, moderate, or severe depression.

#### Cognitive Outcomes

Participants completed three computer-based cognitive tasks (delivered using E-Prime Professional (V2.0, 2012) at age 24 years (*M* = 24.0, *SD =* 0.8). Full descriptions are in the Supplementary Material.

### Emotion Recognition

An emotion recognition task (ERT) assessed accuracy in the recognition of facial displays of six emotions: happiness, sadness, anger, disgust, fear, and surprise. Participants were presented with a series of facial expression and asked to indicate which emotion had been displayed out of six possible emotions. Stimuli ranged in intensity from a near-neutral expression to a prototypical display. The primary outcome measure of the ERT was total hits (the number of correctly identified facial expressions, out of 96). Secondary outcomes included hits by emotion category (out of 16).

### Working Memory

An *n*-back task was used as a continuous performance measure of working memory (Kirchner, 1958). Participants monitored a series of numbers and indicated whether each number matched the one they saw 2 trials previously. Participants responded via keypress. The task consisted of 48 trials, eight of which were target trials (i.e., matches). The primary outcome measure was d prime (*d’*), a discriminability index that takes in to account the proportion of hits (correctly identified matches) to false alarms (non-matches incorrectly identified as matches) to estimate signal-detection ability (McNichol, 1972). Higher *d’* indicates better performance. Participants were excluded if they responded to fewer than 50% of trials or had a negative *d’*.

### Response Inhibition

A stop signal task (Logan et al., 1984) was used to assess participants’ capacity to withhold a motor response. Participants were presented with a series of trials displaying either an “X” or “O” on a blank screen. On each trial, participants were asked to respond by pressing the corresponding key (X or O) as quickly as possible, unless they heard an auditory tone (the “stop signal”). Participants completed 4 blocks of 64 trials, 25% of which had a stop signal. The delay between stimulus onset and stop signal (“stop signal delay”) varied between trials. The primary outcome was stop signal reaction time (SSRT), calculated as the difference between the median reaction time for go trials and an estimate of the median stop signal delay (SSRT = Go Reaction Time_med_ – Median Stop Signal Delay). Median Stop Signal Delay was the latency where each participant was likely to fail to inhibit 50% of trials. Lower SSRTs indicate better response inhibition.

#### Potential Confounders

Adjustment was made for a range of sociodemographic variables previously found to be associated with mental health and cognitive outcomes. These were participant sex, highest parental social class (4 levels using the 1991 Office of Population Census and Statistics Classification (Dale, 1993): unskilled or semi-skilled manual, skilled manual or non-manual, managerial and technical, and professional), mother’s highest education (determined during pregnancy and coded as below O-level, O-level, above O-level, indicating completion of a school-leaving qualification at age 16), housing tenure (owned/mortgaged vs. other), maternal tobacco use during pregnancy (present/absent), mother’s age at birth, and child ethnicity (non-white/white). Confounders relating to cognition were IQ and head injury. IQ was assessed with the Wechsler Abbreviated Scale of Intelligence (vocabulary and matrix reasoning tests; Wechsler, 1999) at age 15, which was the nearest available timepoint to the exposure. Head injury was defined as a cracked skull or loss of consciousness at any timepoint from 0 to 16 years (coded as present/absent), collected via parent- or self-report.

### Statistical Analyses

Primary analyses were conducted in R (R Core Team, 2018).

#### Observational Analyses

We used multivariable linear regression to examine the association between anxiety and depression and the three primary cognitive outcome measures (ERT total hits, SSRT and *d’*). A second multivariable model examined the association between anxiety and depression and individual emotions on the ERT (hit scores for happy, sadness, anger, disgust, fear, and surprise). All models were conducted cross-sectionally (exposure: anxiety and depression assessed at 24 years) and prospectively (exposure: anxiety and depression assessed at 18 years).

Anxiety and depression were initially entered as exposures in separate models. Each model was then adjusted for (i) sociodemographic variables (sex, ethnicity, parental occupation, mother’s education, housing tenure, mother’s age at birth and mother’s tobacco use in pregnancy); (ii) additionally, history of head injury and IQ; and (iii) concurrent anxiety or depression at that timepoint, to determine if there was a unique effect of either mental health exposure. The final model at each timepoint was the same for both anxiety and depression.

We initially conducted complete case analyses, using participants with data for all exposures, outcomes, and confounders (cross-sectional *n* = 2,187; prospective *n* = 1,855). As missing data can lead to biased estimates (Sterne et al., 2009), we conducted multiple imputation using the ‘ice’ package in Stata (StataCorp, 2017). We included a range of auxiliary variables, in addition to all exposure and confounders, to predict missing data and impute 100 datasets. Exposure and confounder data was imputed for all participants who had completed all three cognitive assessments (*n* = 3,087). We present the multiply imputed analyses as the primary results. A comparison of available and missing ALSPAC participants is presented in Supplementary Table S1.

#### Genetic Analyses

We conducted Mendelian randomization (MR) to examine possible causal pathways between cognition and anxiety and depression. We initially sought to conduct MR bidirectionally, using genetic instruments for both cognition and anxiety/depression. However, genome-wide association studies (GWAS) of the three primary cognitive outcomes in ALSPAC did not yield any genome-wide significant single nucleotide polymorphisms (SNPs) to use as genetic instruments (as previously reported by Mahedy et al., 2021). Therefore, we conducted MR analyses in the direction of mental health exposure to cognitive outcome, using published summary statistics from anxiety and depression GWAS as exposures. Analyses were conducted using the TwoSampleMR package 0.4.26 (Hemani et al., 2018).

For depression MR analyses, we used 40 SNPs associated with major depression previously identified by Wray and colleagues (2018) comparing 135,458 cases and 344,901 controls. For anxiety, we used summary statistics from a meta-analysis of GWAS of anxiety disorders performed by Otowa and colleagues (case-control analysis, total *n =* 17,310; Otowa et al., 2016). One genome wide significant SNP was identified. We therefore used a relaxed threshold of *p* < 5×10^−5^, which identified 497 SNPs. These SNPs were clumped at linkage disequilibrium (LD) *r*^2^ = 0.001 and a distance of 10,000kb using the *clump_data* command prior to analysis, resulting in 87 independent loci.

We sought to conduct both one- and two-sample MR. For one-sample, we generated polygenic risk scores (PRS) of anxiety and depression for each participant using PLINK (v. 1.90; Purcell et al., 2007), which summed the number of risk alleles for each SNP weighted by the effect estimate of that SNP in the discovery GWAS. We then ran logistic regressions, regressing each PRS onto anxiety/depression at age 24 in ALSPAC, to confirm that the PRS were internally valid. We then aimed to conduct instrumental variable regressions, regressing the residuals from the PRS to anxiety/depression analyses onto cognitive outcome measures in our ALSPAC sample. For two-sample MR, we compared results across three methods: inverse-variance weighted (IVW), weighted median (Bowden et al., 2016), and weighted mode (Hartwig et al., 2017). IVW is the primary method, and each of the others are sensitivity tests that make different assumptions regarding the validity of the genetic instruments (Bowden et al., 2016; Hartwig et al., 2017). A consistent effect across all methods would provide the most robust evidence for a causal effect.

## Results

### Anxiety and Depression

A breakdown of the sample by anxiety and depression case/control status, alongside summary cognitive scores, can be seen in Supplementary Table S2. At age 18, 7.2% of the complete cases met ICD-10 criteria for a primary or secondary diagnosis of depression, and 8.5% met the criteria for a primary or secondary diagnosis of an anxiety disorder. 3.8% of the sample met criteria for both. At age 24, 9.5% of complete cases met criteria for depression and 12.0% met criteria for an anxiety disorder, while 6.2% met criteria for both anxiety and depression.

### Cross-sectional Analyses

At age 24, there was evidence for a negative association between anxiety and working memory (*d’*, Table 1). This was consistent across all levels of adjustment. There was no clear evidence for an association between depression and working memory. No clear evidence was found for an association between either anxiety or depression and response inhibition (SSRT).

**Table 1.**
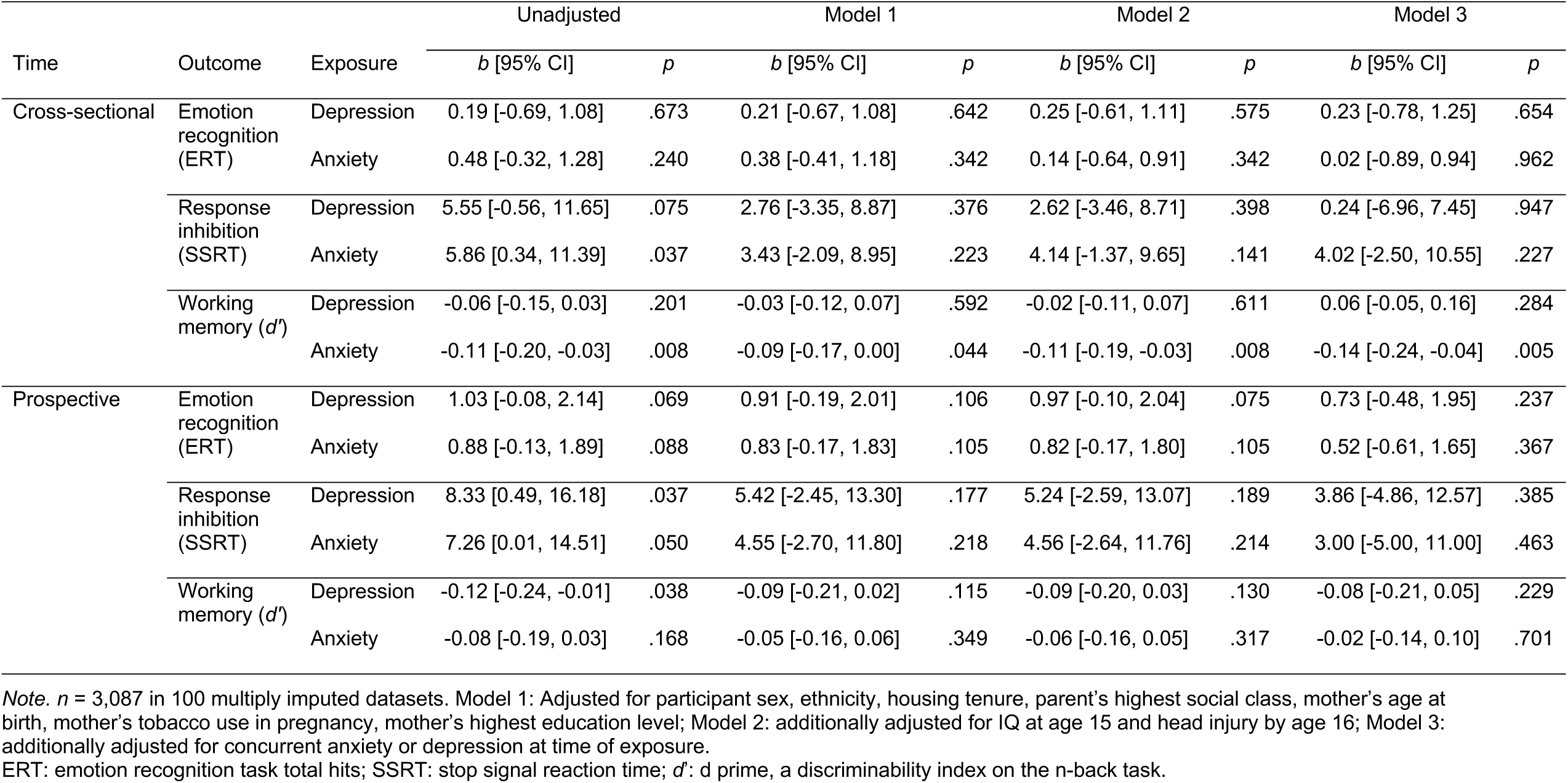
Cross-sectional and prospective associations with emotion recognition, working memory and response inhibition (imputed dataset)

| Time | Outcome | Exposure | Unadjusted |  | Model 1 |  | Model 2 |  | Model 3 |  |
| --- | --- | --- | --- | --- | --- | --- | --- | --- | --- | --- |
|  |  |  | *b* [95% CI] | *p* | *b* [95% CI] | *p* | *b* [95% CI] | *p* | *b* [95% CI] | *p* |
| Cross-sectional | Emotion recognition (ERT) | Depression | 0.19 [-0.69, 1.08] | .673 | 0.21 [-0.67, 1.08] | .642 | 0.25 [-0.61, 1.11] | .575 | 0.23 [-0.78, 1.25] | .654 |
|  |  | Anxiety | 0.48 [-0.32, 1.28] | .240 | 0.38 [-0.41, 1.18] | .342 | 0.14 [-0.64, 0.91] | .342 | 0.02 [-0.89, 0.94] | .962 |
|  | Response inhibition (SSRT) | Depression | 5.55 [-0.56, 11.65] | .075 | 2.76 [-3.35, 8.87] | .376 | 2.62 [-3.46, 8.71] | .398 | 0.24 [-6.96, 7.45] | .947 |
|  |  | Anxiety | 5.86 [0.34, 11.39] | .037 | 3.43 [-2.09, 8.95] | .223 | 4.14 [-1.37, 9.65] | .141 | 4.02 [-2.50, 10.55] | .227 |
|  | Working memory (*d'*) | Depression | -0.06 [-0.15, 0.03] | .201 | -0.03 [-0.12, 0.07] | .592 | -0.02 [-0.11, 0.07] | .611 | 0.06 [-0.05, 0.16] | .284 |
|  |  | Anxiety | -0.11 [-0.20, -0.03] | .008 | -0.09 [-0.17, 0.00] | .044 | -0.11 [-0.19, -0.03] | .008 | -0.14 [-0.24, -0.04] | .005 |
| Prospective | Emotion recognition (ERT) | Depression | 1.03 [-0.08, 2.14] | .069 | 0.91 [-0.19, 2.01] | .106 | 0.97 [-0.10, 2.04] | .075 | 0.73 [-0.48, 1.95] | .237 |
|  |  | Anxiety | 0.88 [-0.13, 1.89] | .088 | 0.83 [-0.17, 1.83] | .105 | 0.82 [-0.17, 1.80] | .105 | 0.52 [-0.61, 1.65] | .367 |
|  | Response inhibition (SSRT) | Depression | 8.33 [0.49, 16.18] | .037 | 5.42 [-2.45, 13.30] | .177 | 5.24 [-2.59, 13.07] | .189 | 3.86 [-4.86, 12.57] | .385 |
|  |  | Anxiety | 7.26 [0.01, 14.51] | .050 | 4.55 [-2.70, 11.80] | .218 | 4.56 [-2.64, 11.76] | .214 | 3.00 [-5.00, 11.00] | .463 |
|  | Working memory (*d'*) | Depression | -0.12 [-0.24, -0.01] | .038 | -0.09 [-0.21, 0.02] | .115 | -0.09 [-0.20, 0.03] | .130 | -0.08 [-0.21, 0.05] | .229 |
|  |  | Anxiety | -0.08 [-0.19, 0.03] | .168 | -0.05 [-0.16, 0.06] | .349 | -0.06 [-0.16, 0.05] | .317 | -0.02 [-0.14, 0.10] | .701 |

*Note.* *n* = 3,087 in 100 multiply imputed datasets. Model 1: Adjusted for participant sex, ethnicity, housing tenure, parent's highest social class, mother's age at birth, mother's tobacco use in pregnancy, mother's highest education level; Model 2: additionally adjusted for IQ at age 15 and head injury by age 16; Model 3: additionally adjusted for concurrent anxiety or depression at time of exposure.
ERT: emotion recognition task total hits; SSRT: stop signal reaction time; *d'*: *d* prime, a discriminability index on the *n*-back task.

There was also no clear evidence for an association between anxiety or depression with global emotion recognition (ERT total hits, Table 1); however, effect estimates were consistently positive. When analysing individual emotions (Table 2), there was some evidence that both anxiety and depression were associated with poorer recognition of happy faces. However, the evidence for the association with anxiety was consistently stronger, and in the final model (including both anxiety and depression), only the anxiety effect remained. In contrast, there was strong evidence for a unique association between depression and an increased recognition of sad faces, which was consistent across all models, where anxiety effects did not survive adjustment for depression. There was no clear evidence for associations with the recognition of any other emotions. Complete case analyses are presented in Supplementary Table S3-S4.

**Table 2.** Cross-sectional associations with emotion-specific hit rate on the Emotion Recognition Task (imputed dataset)

| Emotion | Exposure | Unadjusted |  | Model 1 |  | Model 2 |  | Model 3 |  |
| --- | --- | --- | --- | --- | --- | --- | --- | --- | --- |
|  |  | *b* [95% CI] | *p* | *b* [95% CI] | *p* | *b* [95% CI] | *p* | *b* [95% CI] | *p* |
| Happy | Depression | -0.19 [-0.43, 0.06] | .135 | -0.27 [-0.51, -0.02] | .034 | -0.27 [-0.51, -0.02] | .035 | -0.11 [-0.40, 0.19] | .478 |
|  | Anxiety | -0.22 [-0.45, 0.00] | .049 | -0.32 [-0.54, -0.09] | .005 | -0.32 [-0.55, -0.10] | .005 | -0.27 [-0.54, -0.01] | .045 |
| Sad | Depression | 0.35 [0.11, 0.60] | .005 | 0.38 [0.14, 0.63] | .002 | 0.39 [0.15, 0.63] | .002 | 0.35 [0.07, 0.64] | .016 |
|  | Anxiety | 0.28 [0.06, 0.50] | .014 | 0.29 [0.07, 0.51] | .010 | 0.23 [0.01, 0.45] | .039 | 0.06 [-0.20, 0.32] | .655 |
| Anger | Depression | -0.07 [-0.36, 0.21] | .625 | -0.06 [-0.34, 0.23] | .690 | -0.05 [-0.33, 0.24] | .751 | -0.08 [-0.41, 0.26] | .656 |
|  | Anxiety | 0.07 [-0.19, 0.33] | .588 | 0.08 [-0.18, 0.33] | .568 | 0.01 [-0.24, 0.27] | .914 | 0.05 [-0.25, 0.35] | .741 |
| Disgust | Depression | 0.03 [-0.23, 0.29] | .816 | 0.03 [-0.23, 0.29] | .843 | 0.03 [-0.23, 0.29] | .820 | -0.09 [-0.39, 0.22] | .581 |
|  | Anxiety | 0.21 [-0.02, 0.45] | .075 | 0.19 [-0.04, 0.43] | .109 | 0.16 [-0.08, 0.39] | .194 | 0.20 [-0.08, 0.48] | .164 |
| Surprise | Depression | -0.14 [-0.31, 0.04] | .117 | -0.13 [-0.31, 0.04] | .144 | -0.13 [-0.30, 0.05] | .155 | -0.12 [-0.33, 0.08] | .243 |
|  | Anxiety | -0.06 [-0.22, 0.09] | .425 | -0.06 [-0.22, 0.10] | .453 | -0.07 [-0.23, 0.09] | .416 | -0.01 [-0.19, 0.18] | .951 |
| Fear | Depression | 0.21 [-0.19, 0.60] | .305 | 0.26 [-0.14, 0.65] | .202 | 0.27 [-0.12, 0.66] | .179 | 0.27 [-0.19, 0.73] | .248 |
|  | Anxiety | 0.21 [-0.15, 0.56] | .256 | 0.20 [-0.15, 0.56] | .260 | 0.12 [-0.23, 0.48] | .492 | -0.01 [-0.42, 0.41] | .970 |

*Note.* *n* = 3,087 in 100 multiply imputed datasets. Model 1: Adjusted for participant sex, ethnicity, housing tenure, parent's highest social class, mother's age at birth, mother's tobacco use in pregnancy, mother's highest education level; Model 2: additionally adjusted for IQ at age 15 and head injury by age 16; Model 3: additionally adjusted for concurrent anxiety or depression at time of exposure (age 24).

### Prospective Analyses

There was some evidence that individuals with depression at age 18 performed poorer on both the response inhibition and working memory tasks, and higher on emotion recognition ability (Table 1). However, these associations attenuated after adjustment. There was no clear evidence for a prospective association between anxiety at age 18 and any of the three primary cognitive outcomes.

When analysing individual emotion hits (Supplementary Table S3), there was evidence that both depression and anxiety at age 18 were positively associated with recognition of fearful faces at age 24. These associations were consistent until adjusting for concurrent anxiety and depression. Anxiety at age 18 was also found to have a consistent, positive association with the recognition of disgusted faces at age 24. There was no clear evidence for associations with the recognition of any other emotions.

### Mendelian Randomization

For one-sample MR analyses, neither the anxiety or depression PRS predicted the corresponding phenotype in ALSPAC (anxiety: OR = 1.08, 95% CI = 0.96 to 1.20, *p* = .823; depression: OR = 1.03, 95% CI = 0.91 to 1.17, *p* = .607). Therefore, we were unable to continue this analysis past the validation stage.

Two-sample MR results are presented in Table 4. There was very weak evidence for a possible effect of depression on improved global emotion recognition (IVW estimate: 0.275, 95% CI: -0.04 to 0.56, *p* = .082), with a consistent direction of effect across sensitivity analyses. There was no evidence for causal effects in any of the remaining MR models.

**Table 3.**
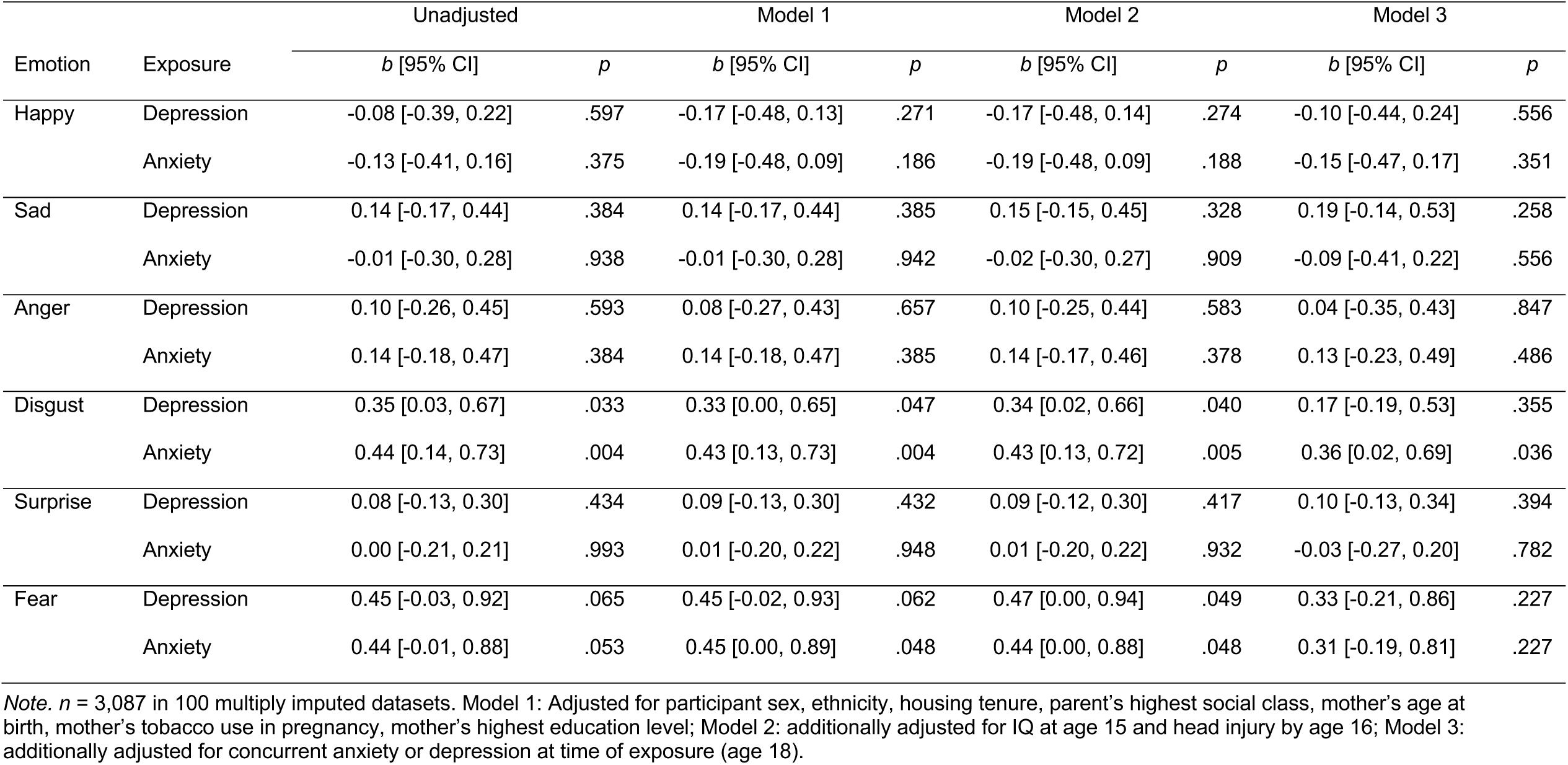
Prospective associations with emotion-specific hit rate on the Emotion Recognition Task (imputed dataset)

**Table 4.** Two sample Mendelian randomization analyses of mental health and cognitive outcomes

| Outcome | Method | Depression<br>(*n* SNPs = 40) |  | Anxiety<br>(*n* SNPs = 72) |  |
| --- | --- | --- | --- | --- | --- |
|  |  | Beta [95%CI] | *p* | Beta [95%CI] | *p* |
| Emotion recognition<br>(ERT total hits) | Inverse-variance weighted | 0.275 [-0.035, 0.585] | .082 | -0.006 [-0.054, 0.042] | .803 |
|  | Weighted median | 0.141 [-0.282, 0.563] | .513 | -0.016 [-0.088, 0.056] | .658 |
|  | Weighted mode | 0.007 [-0.884, 0.897] | .988 | -0.036 [-0.17, 0.098] | .601 |
| Response inhibition<br>(SSRT) | Inverse-variance weighted | 0.014 [-0.332, 0.361] | .935 | -0.010 [-0.058, 0.038] | .682 |
|  | Weighted median | 0.168 [-0.276, 0.613] | .458 | 0.024 [-0.048, 0.096] | .513 |
|  | Weighted mode | 0.429 [-0.465, 1.323] | .353 | 0.053 [-0.086, 0.193] | .457 |
| Working memory (*d'*) | Inverse-variance weighted | 0.082 [-0.213, 0.377] | .586 | -0.027 [-0.075, 0.022] | .282 |
|  | Weighted median | -0.029 [-0.456, 0.398] | .893 | 0.005 [-0.069, 0.079] | .888 |
|  | Weighted mode | -0.650 [-1.602, 0.302] | .189 | 0.081 [-0.084, 0.246] | .341 |

*Note.* ERT: emotion recognition task total hits; SSRT: stop signal reaction time; *d'*: d prime, a discriminability index on the n-back task.

## Discussion

We investigated the relationship of anxiety and depression with three domains of cognition (emotion recognition, working memory and response inhibition) in a population sample of young adults. Cross-sectionally, we observed a negative association between anxiety and working memory performance, as well as both anxiety and depression being associated with differences in emotion recognition accuracy. There was evidence for prospective associations between anxiety and depression at age 18 and emotion recognition at age 24. We sought to triangulate these findings with genetic analyses, however, MR results were inconclusive, due to limited statistical power. This study is one of few cohort studies examining cognition and mental health in young adulthood, a critical time point for the development of disorders (Le Roux et al., 2005; Park et al., 2014). Our results suggest that, while there is some evidence of differences in cognitive functioning, these associations are small.

In terms of executive functioning, there was evidence that anxiety was associated with mild impairments in working memory performance. Previous research has suggested deficits in both anxiety (Ajilchi & Nejati, 2017; Moran, 2016) and depression (Nikolin et al., 2021). There was also no clear evidence that anxiety and depression at age 18 were prospectively associated with either working memory or response inhibition at age 24. While some evidence was found in unadjusted analyses (both cross-sectionally and prospectively), this attenuated when adjusting for key sociodemographic and cognitive variables. Such variables are often unavailable in smaller psychological studies of cognition, highlighting the importance of longitudinal research in this area. Overall, our findings are in line with previous work suggesting that any effect of anxiety and depression on executive functioning is likely to be small (Smitherman et al., 2007).

Emotion recognition ability has received attention as both a potential biomarker and causal mechanism in emotional disorders (Godlewska et al., 2012). Contrary to some previous research (Demenescu et al., 2010), we did not find evidence for a general deficit in global emotion recognition ability. Although the evidence was weak, effect estimates were consistently positive. However, when studying accuracy by emotion, we found strong evidence that both depression and anxiety were associated with a more ‘negative’ pattern of responding. Cross-sectionally, depression was associated with an increased accuracy of recognising sad faces, while anxiety was associated with a decreased accuracy of recognising happy faces. This is consistent with previous research suggesting that depression is associated with a negative bias (Foland-Ross & Gotlib, 2012). A similar pattern was identified prospectively, with both anxiety and depression being associated with superior recognition of fearful faces, and anxiety with disgust. It is unclear why the response pattern varied across timepoints. Taken together, these findings provide support that emotional disorders in young adulthood are associated with aberrant emotional processing.

This study has several limitations. First, it is likely that relationships between cognition and mental health are bidirectional. We were unable to conduct analyses in the direction of cognition to mental health, due to a) no subsequent mental health phenotypic data being available and b) no valid genetic instruments for the cognitive measures studied here. This limited our capacity to assess causal pathways. Nonetheless, this work is an important first step in investigating cognition in ALSPAC, which can be built upon as further clinic data becomes available. To address a lack of genetic instruments for cognition, future work could meta-analyse cognition across cohort studies to improve power. Second, our MR analyses (in the direction of mental health to cognition) also suffered from low power, leading to imprecise estimates, and the depression and anxiety PRS did not predict the corresponding phenotypes in our sample. Lastly, while evidence suggests that anxiety and depression symptoms lie on a continuum (Hankin et al., 2005), we relied on discrete variables available in ALSPAC. This meant we were unable to assess the effect of symptom severity.

There was little evidence that anxiety and depression were associated with significant impairments in executive functioning. However, there were moderate associations with emotion recognition accuracy, with both being associated with increased accuracy in recognising more negative emotions. This may inform the development of interventions that target psychosocial functioning in emotional disorders.

## Data Availability

Any researcher can apply to use ALSPAC data, including the variables under investigation in this study. Access information is provided here: http://www.bristol.ac.uk/alspac/researchers/access/

## Acknowledgements

We are extremely grateful to all the families who took part in this study, the midwives for their help in recruiting them, and the whole ALSPAC team, which includes interviewers, computer and laboratory technicians, clerical workers, research scientists, volunteers, managers, receptionists, and nurses.

## References

Ajilchi, B., & Nejati, V. (2017). Executive functions in students with depression, anxiety, and stress symptoms. Basic and Clinical Neuroscience, 8(3), 223–232. https://doi.org/10.18869/nirp.bcn.8.3.223

Bowden, J., Davey Smith, G., Haycock, P. C., & Burgess, S. (2016). Consistent Estimation in Mendelian Randomization with Some Invalid Instruments Using a Weighted Median Estimator. Genetic Epidemiology, 40(4), 304–314. https://doi.org/10.1002/gepi.21965

Cullen, B., Nicholl, B. I., Mackay, D. F., Martin, D., Ul-Haq, Z., McIntosh, A., Gallacher, J., Deary, I. J., Pell, J. P., Evans, J. J., & Smith, D. J. (2015). Cognitive function and lifetime features of depression and bipolar disorder in a large population sample: Crosssectional study of 143,828 UK Biobank participants. European Psychiatry, 30(8), 950– 958. https://doi.org/10.1016/j.eurpsy.2015.08.006

Dale, A., Creeser, R., Dodgeon, B., Gleave, S., & Filakti, H. (1993). An introduction to the OPCS Longitudinal Study. Environment & Planning A, 25(10), 1387–1398. https://doi.org/10.1068/a251387

Dalili, M. N., Penton-Voak, I. S., Harmer, C. J., & Munafò, M. R. (2015). Meta-analysis of emotion recognition deficits in major depressive disorder. Psychological Medicine, 45(6), 1135–1144. https://doi.org/10.1017/S0033291714002591

De Nooij, L., Harris, M., Adams, M., Clarke, T.-K., Shen, X., Cox, S., McIntosh, A., & Whalley, H. (2020). Cognitive functioning and lifetime Major Depressive Disorder in UK Biobank. European Psychiatry, 63(1), 1–24. https://doi.org/10.1192/j.eurpsy.2020.24

Demenescu, L. R., Kortekaas, R., den Boer, J. A., & Aleman, A. (2010). Impaired Attribution of Emotion to Facial Expressions in Anxiety and Major Depression. PLoS ONE, 5(12), e15058. https://doi.org/10.1371/journal.pone.0015058

E-Prime Professional, V2.0. (2012). Psychology Software Tools, Inc.

Essau, C. A., Lewinsohn, P. M., Lim, J. X., Ho, M. ho R., & Rohde, P. (2018). Incidence, recurrence and comorbidity of anxiety disorders in four major developmental stages. Journal of Affective Disorders, 228, 248–253. https://doi.org/10.1016/j.jad.2017.12.014

Foland-Ross, L. C., & Gotlib, I. H. (2012). Cognitive and Neural Aspects of Information Processing in Major Depressive Disorder: An Integrative Perspective. Frontiers in Psychology, 3, 489. https://doi.org/10.3389/fpsyg.2012.00489

Godlewska, B. R., Norbury, R., Selvaraj, S., Cowen, P. J., & Harmer, C. J. (2012). Shortterm SSRI treatment normalises amygdala hyperactivity in depressed patients. Psychological Medicine, 42(12), 2609–2617. https://doi.org/10.1017/S0033291712000591

Hankin, B. L., Fraley, R. C., Lahey, B. B., & Waldman, I. D. (2005). Is depression best viewed as a continuum or discrete category? A taxometric analysis of childhood and adolescent depression in a population-based sample. Journal of Abnormal Psychology, 114(1), 96–110. https://doi.org/10.1037/0021-843X.114.1.96

Harris, P. A., Taylor, R., Thielke, R., Payne, J., Gonzalez, N., & Conde, J. G. (2009). Research electronic data capture (REDCap)-A metadata-driven methodology and workflow process for providing translational research informatics support. Journal of Biomedical Informatics, 42(2), 377–381. https://doi.org/10.1016/j.jbi.2008.08.010

Hartshorne, J. K., & Germine, L. T. (2015). When Does Cognitive Functioning Peak? The Asynchronous Rise and Fall of Different Cognitive Abilities Across the Life Span. Psychological Science, 26(4), 433–443. https://doi.org/10.1177/0956797614567339

Hartwig, F. P., Smith, G. D., & Bowden, J. (2017). Robust inference in summary data Mendelian randomization via the zero modal pleiotropy assumption. International Journal of Epidemiology, 46(6), 1985–1998. https://doi.org/10.1093/ije/dyx102

Hemani, G., Zheng, J., Elsworth, B., Wade, K. H., Haberland, V., Baird, D., Laurin, C., Burgess, S., Bowden, J., Langdon, R., Tan, V. Y., Yarmolinsky, J., Shihab, H. A., Timpson, N. J., Evans, D. M., Relton, C., Martin, R. M., Davey Smith, G., Gaunt, T. R., & Haycock, P. C. (2018). The MR-base platform supports systematic causal inference across the human phenome. ELife, 7. https://doi.org/10.7554/eLife.34408

Hirschfeld, R. M. A. (2001). The comorbidity of major depression and anxiety disorders: Recognition and management in primary care. Primary Care Companion to the Journal of Clinical Psychiatry, 3(6), 244–254. https://doi.org/10.4088/pcc.v03n0609

Kirchner, W. K. (1958). Age differences in short-term retention of rapidly changing information. Journal of Experimental Psychology, 55(4), 352–358. https://doi.org/10.1037/h0043688

Knight, M. J., Air, T., & Baune, B. T. (2018). The role of cognitive impairment in psychosocial functioning in remitted depression. Journal of Affective Disorders, 235, 129–134. https://doi.org/10.1016/j.jad.2018.04.051

Le Roux, H., Gatz, M., & Wetherell, J. L. (2005). Age at onset of generalized anxiety disorder in older adults. American Journal of Geriatric Psychiatry, 13(1), 23–30. https://doi.org/10.1097/00019442-200501000-00005

Lee, R. S. C., Hermens, D. F., Porter, M. A., & Redoblado-Hodge, M. A. (2012). A metaanalysis of cognitive deficits in first-episode Major Depressive Disorder. In Journal of Affective Disorders (Vol. 140, Issue 2, pp. 113–124). Elsevier. https://doi.org/10.1016/j.jad.2011.10.023

Lewis, G., Pelosi, A. J., Araya, R., & Dunn, G. (1992). Measuring psychiatric disorder in the community: A standardized assessment for use by lay interviewers. Psychological Medicine, 22(2), 465–486. https://doi.org/10.1017/S0033291700030415

Logan, G. D., Cowan, W. B., & Davis, K. A. (1984). On the ability to inhibit simple and choice reaction time responses: A model and a method. Journal of Experimental Psychology: Human Perception and Performance, 10(2), 276–291. https://doi.org/10.1037//00961523.10.2.276

Mahedy, L., Suddell, S., Skirrow, C., Fernandes, G. S., Field, M., Heron, J., Hickman, M., Wootton, R., & Munafò, M. R. (2021). Alcohol use and cognitive functioning in young adults: improving causal inference. Addiction, 116(2), 292–302. https://doi.org/10.1111/add.15100

McNichol, D. (1972). A primer of signal detection theory. Allen and Unwin.

Meijsen, J. J., Campbell, A., Hayward, C., Porteous, D. J., Deary, I. J., Marioni, R. E., & Nicodemus, K. K. (2018). Phenotypic and genetic analysis of cognitive performance in Major Depressive Disorder in the Generation Scotland: Scottish Family Health Study. Translational Psychiatry, 8(1), 63. https://doi.org/10.1038/s41398-018-0111-0

Moran, T. P. (2016). Anxiety and working memory capacity: A meta-analysis and narrative review. Psychological Bulletin, 142(8), 831–864. https://doi.org/10.1037/bul0000051

Nikolin, S., Tan, Y. Y., Schwaab, A., Moffa, A., Loo, C. K., & Martin, D. (2021). An investigation of working memory deficits in depression using the n-back task: A systematic review and meta-analysis. In Journal of Affective Disorders (Vol. 284, pp. 1– 8). Elsevier B.V. https://doi.org/10.1016/j.jad.2021.01.084

O’Toole, M. S., Hougaard, E., & Mennin, D. S. (2013). Social anxiety and emotion knowledge: A meta-analysis. In Journal of Anxiety Disorders (Vol. 27, Issue 1, pp. 98– 108). Pergamon. https://doi.org/10.1016/j.janxdis.2012.09.005

Otowa, T., Hek, K., Lee, M., Byrne, E. M., Mirza, S. S., Nivard, M. G., Bigdeli, T., Aggen, S. H., Adkins, D., Wolen, A., Fanous, A., Keller, M. C., Castelao, E., Kutalik, Z., Der Auwera, S. V., Homuth, G., Nauck, M., Teumer, A., Milaneschi, Y., … Hettema, J. M. (2016). Meta-analysis of genome-wide association studies of anxiety disorders. Molecular Psychiatry, 21(10), 1391–1399. https://doi.org/10.1038/mp.2015.197

Park, S. C., Hahn, S. W., Hwang, T. Y., Kim, J. M., Jun, T. Y., Lee, M. S., Kim, J. B., Yim, H. W., & Park, Y. C. (2014). Does age at onset of first major depressive episode indicate the subtype of major depressive disorder?: The clinical research center for depression study. Yonsei Medical Journal, 55(6), 1712–1720. https://doi.org/10.3349/ymj.2014.55.6.1712

Purcell, S., Neale, B., Todd-Brown, K., Thomas, L., Ferreira, M. A. R., Bender, D., Maller, J., Sklar, P., De Bakker, P. I. W., Daly, M. J., & Sham, P. C. (2007). PLINK: A tool set for whole-genome association and population-based linkage analyses. American Journal of Human Genetics, 81(3), 559–575. https://doi.org/10.1086/519795

R Core Team. (2018). R: A language and environment for statistical computing. R Foundation for Statistical Computing, Vienna, Austria.

Rock, P. L., Roiser, J. P., Riedel, W. J., & Blackwell, A. D. (2014). Cognitive impairment in depression: A systematic review and meta-analysis. Psychological Medicine, 44(10), 2029–2040. https://doi.org/10.1017/S0033291713002535

Schuitevoerder, S., Rosen, J. W., Twamley, E. W., Ayers, C. R., Sones, H., Lohr, J. B., Goetter, E. M., Fonzo, G. A., Holloway, K. J., & Thorp, S. R. (2013). A meta-analysis of cognitive functioning in older adults with PTSD. Journal of Anxiety Disorders, 27(6), 550–558. https://doi.org/10.1016/j.janxdis.2013.01.001

Semkovska, M., Quinlivan, L., O’Grady, T., Johnson, R., Collins, A., O’Connor, J., Knittle, H., Ahern, E., & Gload, T. (2019). Cognitive function following a major depressive episode: a systematic review and meta-analysis. The Lancet Psychiatry, 6(10), 851–861. https://doi.org/10.1016/S2215-0366(19)30291-3

Shin, N. Y., Lee, T. Y., Kim, E., & Kwon, J. S. (2014). Cognitive functioning in obsessivecompulsive disorder: A meta-analysis. In Psychological Medicine (Vol. 44, Issue 6, pp. 1121–1130). Cambridge University Press. https://doi.org/10.1017/S0033291713001803

Smitherman, T. A., Huerkamp, J. K., Miller, B. I., Houle, T. T., & O’jile, J. R. (2007). The relation of depression and anxiety to measures of executive functioning in a mixed psychiatric sample. Archives of Clinical Neuropsychology, 22, 647–654. https://doi.org/10.1016/j.acn.2007.04.007

Snyder, H. R. (2013). Major depressive disorder is associated with broad impairments on neuropsychological measures of executive function: A meta-analysis and review. Psychological Bulletin, 139(1), 81–132. https://doi.org/10.1037/a0028727

StataCorp. (2017). Stata Statistical Software: Release 15. StataCorp LLC.

Sterne, J. A. C., White, I. R., Carlin, J. B., Spratt, M., Royston, P., Kenward, M. G., Wood, A. M., & Carpenter, J. R. (2009). Multiple imputation for missing data in epidemiological and clinical research: Potential and pitfalls. In BMJ (Online) (Vol. 339, Issue 7713, pp. 157–160). BMJ. https://doi.org/10.1136/bmj.b2393

Wechsler, D. (1999). Wechsler Abbreviated Scale of Intelligence. The Psychological Corporation: Harcourt Brace & Company.

World Health Organization. (2017). Depression and Other Common Mental Disorders Global Health Estimates.

Wray, N. R., Ripke, S., Mattheisen, M., Trzaskowski, M., Byrne, E. M., Abdellaoui, A., Adams, M. J., Agerbo, E., Air, T. M., Andlauer, T. M. F., Bacanu, S.-A., Bækvad-Hansen, M., Beekman, A. F. T., Bigdeli, T. B., Binder, E. B., Blackwood, D. R. H., Bryois, J., Buttenschøn, H. N., Bybjerg-Grauholm, J., … Major Depressive Disorder Working Group of the Psychiatric Genomics Consortium. (2018). Genome-wide association analyses identify 44 risk variants and refine the genetic architecture of major depression. Nature Genetics, 50(5), 668–681. https://doi.org/10.1038/s41588018-0090-3

