## Supplementary Material for "Cognitive functioning in anxiety and depression: Results from the ALSPAC cohort"

**Supplementary Materials**

**Supplementary Table S1.** Comparison of sociodemographic characteristics of available and unavailable participants (complete case)

| | Available (n = 2187) | Not available (*n* = 12454) | |
|---|---|---|---|
| | *n* (%) | *n* (%) | OR [95% CI] |
| Sex (male) | 865 (40) | 6448 (54) | 0.56 [0.51, 0.61] |
| Ethnicity (white) | 2117 (97) | 9415 (95) | 1.74 [1.36, 2.27] |
| Parent's highest social class (i. unskilled) | 126 (6) | 251 (3) | Ref |
| ii. skilled manual or non-manual | 685 (31) | 1949 (21) | 0.70 [0.56, 0.88] |
| iii. managerial and technical | 1093 (50) | 5146 (55) | 0.42 [0.34, 0.53] |
| iv. professional | 283 (13) | 2034 (22) | 0.28 [0.22, 0.36] |
| Housing tenure (owned/mortgaged) | 1955 (89) | 7925 (70) | 3.60 [3.13, 4.16] |
| Mother's tobacco use (yes) | 274 (13) | 3090 (28) | 0.37 [0.33, 0.43] |
| Mother's education (< O level) | 287 (13) | 3468 (34) | Ref |
| O level | 715 (32.7) | 3611 (35) | 2.39 [2.07, 2.77] |
| > O level | 1185 (54.18) | 3222 (31) | 4.44 [3.87, 5.10] |
| Head injury (yes) | 192 (9) | 585 (7) | 1.35 [1.14, 1.60] |
| | *M (SD)* | *M (SD)* | *b* [95 CI] |
| Mother's age | 29.8 (5.0) | 27.7 (5.0) | 2.16 [1.94, 2.38] |
| IQ at 15 | 97.9 (12.6) | 91.6 (12.8) | 6.24 [5.53, 6.94] |

*Note.* Mother's education: <O level indicating no qualification; O level: indicating completion of school examinations at age 16; and >O level: indicating completion of college or university education at or after age 18. Head injury: any loss of consciousness or broken skull between the ages of 0 and 16 years.

**Supplementary Table S2.** Summary of complete case sample by anxiety and depression status.

| | | | Depression | | Anxiety | |
|---|---|---|---|---|---|---|
| Analysis | Variable | All (%) | Case (%) | Control (%) | Case (%) | Control (%) |
| Prospective | *n* | 1855 | 136 | 1719 | 160 | 1695 |
| | Female | 1130 (61) | 102 (75) | 1028 (60) | 118 (74) | 1012 (60) |
| | Ethnicity (white) | 1794 (97) | 133 (98) | 1661 (97) | 156 (98) | 1638 (97) |
| | Parent's highest social class (unskilled) | 235 (13) | 19 (14) | 216 (13) | 27 (17) | 208 (12) |
| | ii. skilled manual or non-manual | 937 (51) | 68 (50) | 869 (51) | 77 (48) | 860 (51) |
| | iii. managerial and technical | 573 (31) | 40 (29) | 533 (31) | 44 (28) | 529 (31) |
| | iv. professional | 110 (6) | 9 (7) | 101 (6) | 12 (8) | 98 (6) |
| | Housing tenure (owned/mortgaged) | 1664 (90) | 113 (83) | 1551 (90) | 134 (84) | 1530 (90) |
| | Head injury (yes) | 162 (9) | 11 (8) | 151 (9) | 19 (12) | 143 (8) |
| | IQ* | 98.4 (12.5) | 97.8 (11.9) | 98.5 (12.5) | 98.0 (12.5) | 98.5 (12.5) |
| | Response Inhibition (SSRT)* | 252.9 (50.8) | 260.8 (53.8) | 252.3 (50.6) | 256.4 (54.5) | 252.6 (50.5) |
| | Working Memory (*d'*)* | 2.82 (0.8) | 2.7 (0.8) | 2.8 (0.8) | 2.7 (0.9) | 2.83 (0.8) |
| | ERT total hits* | 67 (7.6) | 68.2 (6.1) | 66.9 (7.6) | 68.1 (7.2) | 66.9 (7.6) |
| Cross-sectional | *n* | 2187 | 208 | 1979 | 262 | 1925 |
| | Female | 1322 (60) | 140 (67) | 1182 (60) | 192 (73) | 1130 (59) |
| | Ethnicity (white) | 2117 (97) | 199 (96) | 1918 (97) | 254 (97) | 1863 (97) |
| | Parent's highest social class (unskilled) | 283 (13) | 33 (16) | 250 (13) | 31 (12) | 252 (13) |
| | ii skilled manual or non-manual | 1093 (50) | 101 (49) | 992 (50) | 137 (52) | 956 (50) |
| | iii. managerial and technical | 685 (31) | 58 (28) | 627 (32) | 74 (28) | 611 (32) |
| | iv. professional | 126 (6) | 16 (8) | 110 (6) | 20 (8) | 106 (6) |
| | Housing tenure (owned/mortgaged) | 1955 (89) | 174 (84) | 1781 (90) | 216 (82) | 1739 (90) |
| | Head injury (yes) | 192 (9) | 23 (11) | 169 (9) | 34 (13) | 158 (8) |
| | IQ at 15* | 97.9 (12.6) | 100.0 (12.7) | 97.9 (12.5) | 99.4 (12.6) | 97.7 (12.5) |
| | Response Inhibition (SSRT)* | 254.2 (51.5) | 258.9 (54.8) | 253.7 (51.5) | 258 (52.7) | 253.6 (51.3) |
| | Working Memory (*d'*)* | 2.8 (0.8) | 2.8 (0.7) | 2.8 (0.8) | 2.8 (0.8) | 2.8 (0.8) |
| | ERT total hits* | 66.9 (7.6) | 67.3 (7.7) | 66.8 (7.6) | 67.4 (7.3) | 66.8 (7.6) |

*Note.* Mother’s education: <O level indicating no qualification; O level: indicating completion of school examinations at age 16; and >O level: indicating completion of college or university education at or after age 18. Head injury: any loss of consciousness or broken skull between the ages of 0 and 16 years; SSRT: stop signal reaction time; ERT: emotion recognition task.
Anxiety and depression cases overlap by *n* = 70 at age 18, and *n* = 136 at age 24.
*Mean (standard deviation).

**Supplementary Table S3.** Cross-sectional and prospective associations with emotion recognition, working memory and response inhibition (complete cases)

| | | | Unadjusted | | Model 1 | | Model 2 | | Model 3 | |
|---|---|---|---|---|---|---|---|---|---|---|
| Time | Outcome | Exposure | *b* [95% CI] | *p* | *b* [95% CI] | *p* | *b* [95% CI] | *p* | *b* [95% CI] | *p* |
| Cross-sectional (*n* = 2,187) | Emotion recognition (ERT) | Depression | 0.40 [-0.69, 1.49] | .470 | 0.32 [-0.75, 1.40] | .555 | 0.25 [-0.80, 1.29] | .645 | 0.34 [-0.89, 1.57] | .589 |
| | | Anxiety | 0.53 [-0.45, 1.52] | .287 | 0.31 [-0.67, 1.28] | .537 | 0.00 [-0.95, 0.95] | .998 | -0.16 [-1.28, 0.96] | .778 |
| | Response inhibition (SSRT) | Depression | 5.22 [-2.14, 12.57] | .164 | 3.94 [-3.42, 11.31] | .294 | 4.11 [-3.23, 11.44] | .272 | 3.08 [-5.55, 11.7] | .484 |
| | | Anxiety | 4.36 [-2.28, 11.00] | .198 | 2.48 [-4.20, 9.16] | .467 | 3.25 [-3.42, 9.93] | .339 | 1.78 [-6.07, 9.63] | .656 |
| | Working memory (*d′*) | Depression | 0.03 [-0.08, 0.14] | .584 | 0.05 [-0.06, 0.16] | .354 | 0.04 [-0.06, 0.15] | .424 | 0.11 [-0.01, 0.24] | .084 |
| | | Anxiety | -0.06 [-0.16, 0.04] | .240 | -0.03 [-0.13, 0.07] | .502 | -0.06 [-0.16, 0.03] | .202 | -0.12 [-0.23, 0.00] | .046 |
| Prospective (*n* = 1,855) | Emotion recognition (ERT) | Depression | 1.28 [-0.03, 2.60] | .057 | 1.15 [-0.16, 2.45] | .085 | 1.12 [-0.14, 2.38] | .083 | 0.81 [-0.59, 2.20] | .256 |
| | | Anxiety | 1.19 [-0.04, 2.41] | .058 | 1.00 [-0.21, 2.21] | .107 | 1.00 [-0.17, 2.18] | .095 | 0.68 [-0.61, 1.98] | .300 |
| | Response inhibition (SSRT) | Depression | 8.49 [-0.38, 17.36] | .061 | 6.61 [-2.28, 15.50] | .145 | 6.69 [-2.17, 15.54] | .139 | 6.96 [-2.81, 16.73] | .163 |
| | | Anxiety | 3.86 [-4.38, 12.10] | .358 | 2.19 [-6.07, 10.45] | .603 | 2.12 [-6.12, 10.35] | .614 | -0.61 [-9.69, 8.47] | .895 |
| | Working memory (*d′*) | Depression | -0.12 [-0.26, 0.01] | .076 | -0.09 [-0.23, 0.04] | .171 | -0.10 [-0.23, 0.04] | .152 | -0.08 [-0.22, 0.07] | .293 |
| | | Anxiety | -0.09 [-0.21, 0.04] | .174 | -0.07 [-0.19, 0.06] | .278 | -0.07 [-0.19, 0.05] | .254 | -0.04 [-0.18, 0.09] | .554 |

*Note.* Model 1: Adjusted for participant sex, ethnicity, housing tenure, parent's highest social class, mother's age at birth, mother's tobacco use in pregnancy, mother's highest education level; Model 2: additionally adjusted for IQ at age 15 and head injury by age 16; Model 3: additionally adjusted for concurrent anxiety or depression at time of exposure.

ERT: emotion recognition task – total hits; SSRT: stop signal reaction time.

**Supplementary Table S4.** Cross-sectional associations with emotion-specific hit rate on the Emotion Recognition Task (complete cases)

| | | Unadjusted | | Model 1 | | Model 2 | | Model 3 | |
|---|---|---|---|---|---|---|---|---|---|
| Emotion | Exposure | *b* [95% CI] | *p* | *b* [95% CI] | *p* | *b* [95% CI] | *p* | *b* [95% CI] | *p* |
| Happy | Depression | -0.33 [-0.64, -0.02] | .037 | -0.39 [-0.69, -0.08] | .014 | -0.39 [-0.70, -0.08] | .014 | -0.14 [-0.50, 0.22] | .451 |
| | Anxiety | -0.37 [-0.65, -0.10] | .008 | -0.49 [-0.77, -0.21] | .001 | -0.50 [-0.78, -0.22] | .001 | -0.43 [-0.76, -0.10] | .010 |
| Sad | Depression | 0.53 [0.22, 0.83] | .001 | 0.55 [0.25, 0.85] | .000 | 0.53 [0.23, 0.82] | .001 | 0.52 [0.17, 0.87] | .004 |
| | Anxiety | 0.33 [0.06, 0.61] | .018 | 0.34 [0.06, 0.61] | .016 | 0.26 [-0.01, 0.53] | .061 | 0.01 [-0.31, 0.33] | .953 |
| Anger | Depression | -0.03 [-0.38, 0.32] | .856 | -0.03 [-0.38, 0.32] | .856 | -0.05 [-0.39, 0.29] | .765 | -0.05 [-0.45, 0.35] | .801 |
| | Anxiety | 0.07 [-0.25, 0.38] | .674 | 0.05 [-0.26, 0.37] | .735 | -0.03 [-0.34, 0.29] | .872 | 0.00 [-0.37, 0.37] | .997 |
| Disgust | Depression | 0.04 [-0.28, 0.36] | .814 | 0.01 [-0.31, 0.33] | .954 | 0.00 [-0.32, 0.31] | .981 | -0.08 [-0.45, 0.29] | .675 |
| | Anxiety | 0.19 [-0.10, 0.47] | .202 | 0.14 [-0.15, 0.43] | .337 | 0.09 [-0.20, 0.38] | .527 | 0.13 [-0.21, 0.47] | .448 |
| Surprise | Depression | -0.13 [-0.34, 0.09] | .251 | -0.13 [-0.35, 0.08] | .230 | -0.13 [-0.35, 0.09] | .235 | -0.15 [-0.40, 0.11] | .258 |
| | Anxiety | -0.03 [-0.22, 0.16] | .758 | -0.04 [-0.24, 0.16] | .685 | -0.04 [-0.24, 0.15] | .669 | 0.03 [-0.20, 0.26] | .817 |
| Fear | Depression | 0.32 [-0.16, 0.80] | .190 | 0.32 [-0.16, 0.79] | .198 | 0.29 [-0.18, 0.77] | .224 | 0.23 [-0.32, 0.79] | .409 |
| | Anxiety | 0.35 [-0.08, 0.79] | .112 | 0.30 [-0.13, 0.74] | .175 | 0.22 [-0.22, 0.65] | .327 | 0.10 [-0.40, 0.61] | .690 |

*Note.* $n$ = 2,187. Model 1: Adjusted for participant sex, ethnicity, housing tenure, parent's highest social class, mother's age at birth, mother's tobacco use in pregnancy, mother's highest education level; Model 2: additionally adjusted for IQ at age 15 and head injury by age 16; Model 3: additionally adjusted for concurrent anxiety or depression at time of exposure (age 24).

**Supplementary Table S5.** Prospective associations with emotion-specific hit rate on the Emotion Recognition Task (complete cases)

| | | Unadjusted | | Model 1 | | Model 2 | | Model 3 | |
|---|---|---|---|---|---|---|---|---|---|
| Emotion | Exposure | *b* [95% CI] | *p* | *b* [95% CI] | *p* | *b* [95% CI] | *p* | *b* [95% CI] | *p* |
| Happy | Depression | -0.18 [-0.55, 0.20] | .362 | -0.28 [-0.66, 0.09] | .141 | -0.28 [-0.66, 0.09] | .140 | -0.21 [-0.63, 0.20] | .312 |
| | Anxiety | -0.15 [-0.50, 0.19] | .386 | -0.24 [-0.59, 0.11] | .182 | -0.24 [-0.59, 0.11] | .185 | -0.15 [-0.54, 0.23] | .437 |
| Sad | Depression | 0.16 [-0.21, 0.53] | .399 | 0.16 [-0.21, 0.53] | .388 | 0.16 [-0.21, 0.52] | .395 | 0.21 [-0.19, 0.61] | .294 |
| | Anxiety | -0.01 [-0.36, 0.33] | .934 | -0.03 [-0.38, 0.31] | .845 | -0.04 [-0.38, 0.30] | .809 | -0.13 [-0.50, 0.25] | .508 |
| Anger | Depression | 0.06 [-0.37, 0.49] | .781 | 0.08 [-0.35, 0.51] | .713 | 0.07 [-0.34, 0.49] | .733 | 0.02 [-0.44, 0.48] | .923 |
| | Anxiety | 0.12 [-0.27, 0.52] | .539 | 0.12 [-0.28, 0.51] | .555 | 0.12 [-0.27, 0.51] | .548 | 0.11 [-0.32, 0.54] | .615 |
| Disgust | Depression | 0.42 [0.03, 0.81] | .035 | 0.37 [-0.02, 0.76] | .060 | 0.37 [-0.02, 0.76] | .061 | 0.18 [-0.25, 0.60] | .419 |
| | Anxiety | 0.54 [0.18, 0.09] | .003 | 0.49 [0.13, 0.85] | .008 | 0.50 [0.14, 0.85] | .007 | 0.43 [0.03, 0.82] | .035 |
| Surprise | Depression | 0.16 [-0.10, 0.42] | .227 | 0.15 [-0.11, 0.41] | .249 | 0.15 [-0.11, 0.41] | .252 | 0.17 [-0.11, 0.46] | .235 |
| | Anxiety | 0.02 [-0.22, 0.26] | .865 | 0.02 [-0.22, 0.26] | .878 | 0.02 [-0.22, 0.26] | .868 | -0.05 [-0.31, 0.22] | .726 |
| Fear | Depression | 0.66 [0.07, 1.25] | .028 | 0.66 [0.07, 1.25] | .028 | 0.65 [0.07, 1.23] | .028 | 0.44 [-0.20, 1.07] | .181 |
| | Anxiety | 0.67 [0.13, 1.22] | .016 | 0.64 [0.09, 1.18] | .021 | 0.65 [0.11, 1.18] | .019 | 0.48 [-0.12, 1.07] | .116 |

*Note.* $n$ = 1,855. Model 1: Adjusted for participant sex, ethnicity, housing tenure, parent's highest social class, mother's age at birth, mother's tobacco use in pregnancy, mother's highest education level; Model 2: additionally adjusted for IQ at age 15 and head injury by age 16; Model 3: additionally adjusted for concurrent anxiety or depression at time of exposure (age 18).

**Full description of cognitive outcomes**

**Emotion Recognition.** An emotion recognition task (ERT) assessed accuracy in the recognition of facial displays of six emotions: happiness, sadness, anger, disgust, fear and surprise. Participants were presented with a series of facial expressions and asked to indicate which emotion had been displayed in a six alternative forced choice design. Each image was displayed for 200ms and was immediately followed by a backwards mask of visual noise (250ms) to prevent processing of afterimages. The six emotion descriptors were then displayed until the participant made a choice, via mouse click. The stimulus set consisted of both male and female Caucasian faces, with 8 levels of intensity per emotion, ranging from a near-neutral expression to a prototypical display (details of how stimuli were created are reported by Attwood and colleagues (2017). Each individual stimulus was presented twice, resulting in a total of 96 trials. The primary outcome measure of the ERT was total hits (the number of correctly identified facial expressions), out of 96. Secondary outcomes included number of hits by emotion (i.e., the number of times an emotion is chosen incorrectly) Hits were out of 16, with higher scores indexing better emotion recognition.

**Working Memory.** An *n*-back task was used as a continuous performance task of working memory (Kirchner, 1958). Participants monitored a series of numbers and were asked to indicate whether each number matched the one they saw 2 trials previously. Each trial presented a number (0-9) on a blank screen for 500ms, immediately followed by a 3000ms response window. Participants responded via keypress, pressing "1" if it matched or "2" if it did not. The task consisted of 48 trials, eight of which were target trials (i.e., matches). The primary outcome measure was d prime (*d'*), a discriminability index that takes in to account the proportion of hits (correctly identified matches) to false alarms (non-matches incorrectly identified as matches) to estimate signal-detection ability (McNichol, 1972). A higher *d'* indicates better working memory performance. Participants were excluded if they responded to fewer than 50% of trials or had a negative *d'*.

**Response Inhibition.** A stop signal task (Logan et al., 1984) was used to assess participants' capacity to withhold a motor response. Participants were presented with a series of trials displaying either an "X" or "O" on a blank screen. On each trial, participants were asked to respond by pressing the corresponding key (X or O) as quickly as possible, unless they heard an auditory tone ("stop signal") indicating they should withhold their response. Participants completed 32 practice trials, followed by 4 experimental blocks of 64 trials, 25% of which had a stop signal. The delay between stimulus onset and stop signal was drawn from one of four adaptive staircases at 100ms, 200ms, 400ms or 500ms. On successful inhibition the staircase was adjusted by -25ms, and on failed inhibition it was adjusted by +25 (possible range: 25ms to 800ms). The primary outcome was stop signal reaction time (SSRT), calculated as the difference between the median reaction time for go trials and an estimate of the median stop signal delay (SSRT = Go Reaction $\text{Time}_{med}$ – Median Stop Signal Delay). Median Stop Signal Delay was the latency where each participant was likely to fail to inhibit 50% of trials. Lower SSRTs indicate better response inhibition.

### References


Attwood, A. S., Easey, K. E., Dalili, M. N., Skinner, A. L., Woods, A., Crick, L., Ilett, E., Penton-Voak, I. S., & Munafò, M. R. (2017). State anxiety and emotional face recognition in healthy volunteers. *Royal Society Open Science*, *4*(5), 160855. https://doi.org/10.1098/rsos.160855

Kirchner, W. K. (1958). Age differences in short-term retention of rapidly changing information. *Journal of Experimental Psychology*, *55*(4), 352–358. https://doi.org/10.1037/h0043688

Logan, G. D., Cowan, W. B., & Davis, K. A. (1984). On the ability to inhibit simple and choice reaction time responses: A model and a method. *Journal of Experimental Psychology: Human Perception and Performance*, *10*(2), 276–291. https://doi.org/10.1037//0096-1523.10.2.276

McNichol, D. (1972). *A primer of signal detection theory*. Allen and Unwin.
